# Do dietary habits, dietary patterns, and macronutrient intakes affect aging? Multi-dimensional evidence from UK Biobank

**DOI:** 10.1101/2023.09.12.23295459

**Authors:** Chen Zhu, Youfa Wang, Xiaosong Yang, Qiran Zhao, Wenyan Xu, Xiaolu Wang, Yanjun Liang, Qihui Chen, Shenggen Fan

## Abstract

The role of diet in aging is pivotal, yet existing research offers inconsistent findings regarding the impact of specific diets on human aging. We conducted a systematic investigation into the relationship between dietary factors and aging, exploring potential causal links between macronutrient intake and aging. Utilizing data from the UK Biobank baseline survey and a 24-hour dietary assessment survey, we employed a High-dimensional Fixed Effects (HDFE) model to examine dietary factors’ association with aging. Multivariable Mendelian Randomization (MVMR) and Semiparametric Nonlinear Mendelian Randomization (NLMR) techniques assessed causal links between macronutrient consumption and aging. HDFE analysis indicated that a healthier diet was generally linked to better aging outcomes, with various dietary components correlating with aging. For instance, plant-based food intake was associated with increased telomere length and/or reduced phenotypic age, while animal-based food consumption correlated with adverse aging effects. MVMR revealed the benefits of carbohydrate intake on aging, reducing phenotypic age (βC=C-0.0025; 95% CI=[-0.0047, -0.0003]; *p* = 0.026) and increasing whole-brain grey matter volume (βC=C0.0262; 95% CI=[0.007, 0.046]; *p* = 0.008). Overall, our study underscores diet’s significant role in biological aging, highlighting the potential advantages of a carbohydrate-rich diet in promoting healthy aging.

## Introduction

Aging is a complex biological process that involves molecular changes, cellular senescence, and physiological dysregulation.^[1]^ Contrary to chronological age, which increases at the same rate for all people, biological aging can progress at different rates in different people and across circumstances.^[2]^ According to geroscience theory, interventions to slow down biological aging can prevent disease, prolong healthy life, and reduce the socioeconomic burden of disease.^[1,3]^ Recent research has also documented significant associations between aging and various lifestyle factors, such as alcohol drinking, smoking, physical activity, and childhood adversity.^[4–6]^

Dietary factors are also found to be linked with many age-related outcomes, including chronic diseases, serum biomarkers, and DNA methylation, suggesting a role a healthy diet may play in delaying aging.^[7–9]^ However, due to diverse measures of biological aging, limited sample sizes, and lack of randomized controlled trials (RCTs), existing findings are mostly inconclusive regarding the effects of specific dietary patterns or consumption of individual food items.^[10]^ For example, while some studies reported that a low-carbohydrate diet led to improvements in serum factors related to the aging process,^[11]^ others found associations between a low-carbohydrate diet and increased mortality.^[12]^ Recent reviews on the effects of diet on cognitive aging and telomere length also reported inconsistent findings.^[13,14]^

The current study aims to systemically investigate the associations between dietary factors and multi-dimensional measures of aging, in particular, the causal effects of macronutrient intakes on aging using cohort study data.

## Results

### Summary statistics

Among all 493,478 participants, 45.7% are male; 54.3% are female (Table 1). Over 90% are White; 32.2% had a college/university degree. The average participant was 56.5 years old, with a BMI of 27.2 kg/m^2^ and a TDI of -1.3, consuming 255 grams of carbohydrates, 78 grams of fat, 82 grams of protein, and a total of 2,121 calories daily.

**Table 1.** Summary statistics of the analytical sample (N = 493,478)

|  |  | Pooled<br>N = 493,478<br>(100%) |  | Male<br>N = 225,676<br>(45.7%) |  | Female<br>N = 267,802<br>(54.3%) |  |
| --- | --- | --- | --- | --- | --- | --- | --- |
| Variables |  | % or<br>Mean | SD. | % or<br>Mean | SD. | % or<br>Mean | SD. |
| Age |  | 56.5 | 8.1 | 56.8 | 8.2 | 56.4 | 8.0 |
| College |  | 32.2% | - | 33.6% | - | 31.1% | - |
| BMI (kg/m<sup>2</sup>) |  | 27.2 | 4.4 | 27.7 | 4.0 | 26.8 | 4.7 |
| Overweight |  | 66.6% | - | 74.6% | - | 59.8% | - |
| Obesity |  | 23.6% | - | 24.9% | - | 22.5% | - |
| Townsend Deprivation Index<sup>a</sup> |  | -1.3 | 3.1 | -1.3 | 3.1 | -1.4 | 3.0 |
| Daily intake of calories |  | 2120.9 | 642.7 | 2300.1 | 675.3 | 1974.1 | 574.5 |
| Daily intake of carbohydrates (grams) |  | 254.9 | 89.5 | 273.1 | 93.3 | 240.0 | 83.4 |
| Daily intake of fat (grams) |  | 77.9 | 30.6 | 83.8 | 32.7 | 73.0 | 27.9 |
| Daily intake of protein (grams) |  | 82.4 | 26.0 | 87.3 | 27.8 | 78.3 | 23.6 |
| Ethnicity | *White* | 91.4% | - | 91.7% | - | 91.2% | - |
|  | *African* | 1.4% | - | 1.3% | - | 1.5% | - |
|  | *East Asian* | 0.4% | - | 0.3% | - | 0.4% | - |
|  | *South Asian* | 1.7% | - | 2.0% | - | 1.4% | - |
|  | *Other<sup>b</sup>* | 5.1% | - | 4.7% | - | 5.5% | - |
| Region | *East Midlands* | 8.0% | - | 8.0% | - | 8.0% | - |
|  | *East of England* | 0.3% | - | 0.3% | - | 0.3% | - |
|  | *London* | 12.4% | - | 12.0% | - | 12.7% | - |
|  | *North East* | 11.1% | - | 11.2% | - | 11.1% | - |
|  | *North West* | 16.3% | - | 16.7% | - | 16.0% | - |
|  | *Scotland* | 7.3% | - | 7.1% | - | 7.4% | - |
|  | *South East* | 9.8% | - | 9.6% | - | 10.0% | - |
|  | *South West* | 8.4% | - | 8.1% | - | 8.7% | - |
|  | *Wales* | 4.3% | - | 4.3% | - | 4.3% | - |
|  | *West Midlands* | 8.2% | - | 8.8% | - | 7.7% | - |
|  | *Yorkshire and The Humber* | 14.0% | - | 14.0% | - | 14.0% | - |

Note: <sup>a</sup> The Townsend Deprivation Index is a measure of local material deprivation in UK Biobank, in which positive numbers indicate lower socioeconomic status, and negative numbers indicate higher socioeconomic status. <sup>b</sup> Other includes any races or ethnicities not otherwise specified.

### Associations of dietary factors with aging

Table 2, panel A, presents HDFE estimates of equation (1), revealing statistically significant associations between aging and several food/beverage intakes. Animal-based foods tend to be adversely related to aging. For example, processed meat intake is associated with reduced telomere length (Model 1), increased phenotypic age (Model 2), and reduced brain grey matter volume (Model 3). Similarly, poultry, beef, lamb, and pork intakes are linked with reduced telomere length and/or increased phenotypic age.^1^ Intakes of oily fish, processed meat, lamb, and cheese are associated with reduced brain grey/white volumes. In contrast, plant-based foods seem to have potential anti-aging effects. Intakes of plant-based foods (cooked/raw vegetables, fresh/dried fruits, and cereal) are associated with increased telomere length and/or reduced phenotypic age. Cereal intake is also associated with longer telomere length (Model 1) and younger phenotypic age (Model 2). Bread and cereal intakes seem to have a protective effect on brain grey/white matter volumes (Models 3-4).

**Table 2.**
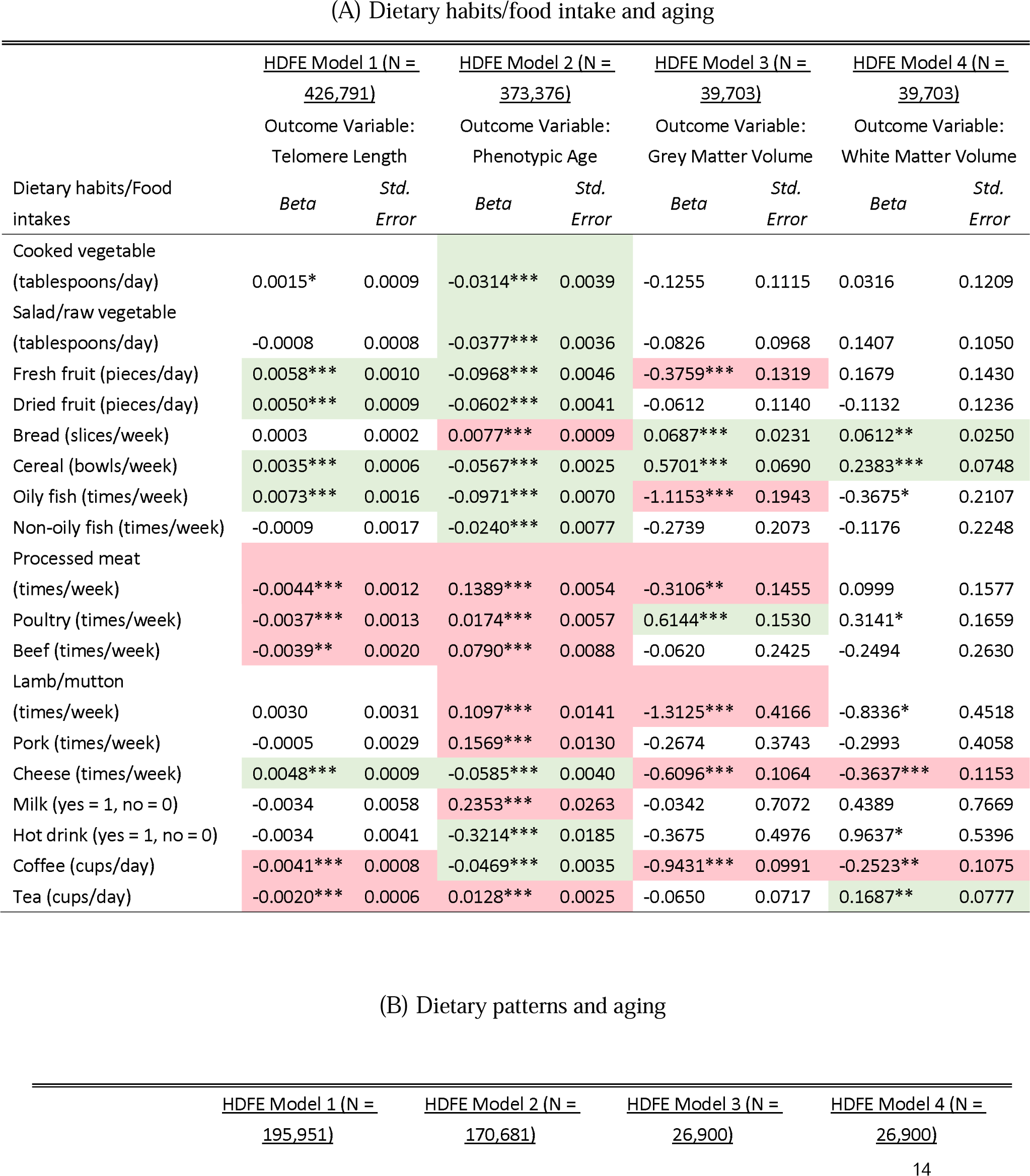
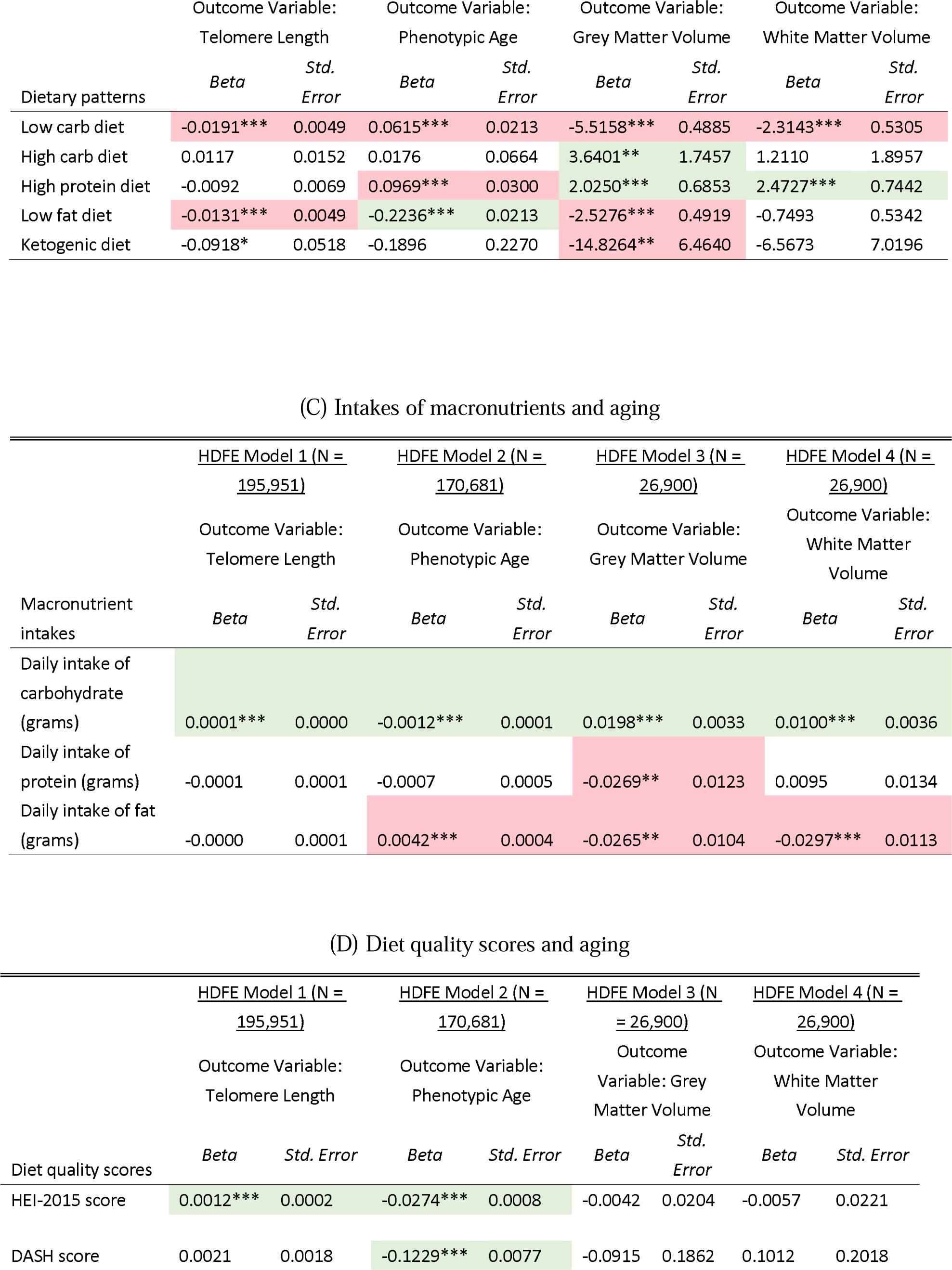
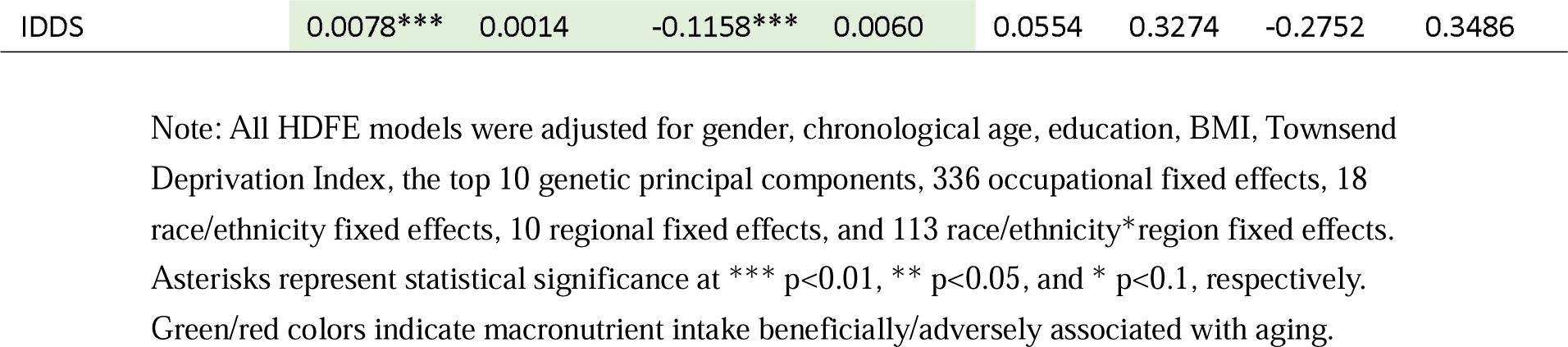
Associations of dietary habits, dietary patterns, macronutrient intakes, and diet quality scores with different measures of aging from high-dimensional fixed effects estimation.

The results on some other food/beverage intakes are mixed. For instance, fresh fruit intake is beneficially associated with longer telomere length (Model 1) and younger phenotypic age (Model 2) but adversely associated with reduced grey matter volume (Model 3). Poultry intake is linked with reduced telomere length and increased phenotypic age but with increased brain grey/white matter volumes. Surprisingly, coffee is adversely associated with decreased telomere length and grey/white matter volume, though beneficially with reduced phenotypic age. Tea is adversely associated with decreased telomere length and increased phenotypic age but beneficially with increased white matter volume. Hot drinks are associated with younger phenotypic age.

Table 2, panel B, reports estimated associations of certain dietary patterns/macronutrient intakes with aging. Surprisingly, the low-carbohydrate diet (defined as any diet containing carbohydrates <45% of daily total energy intake^[32]^) is adversely associated with all four measures of aging (Models 1-4). For grey matter volume, the high-carbohydrate diet (defined as any diet containing carbohydrates of >65% of daily total energy intake^[32,33]^) shows a positive/beneficial link, while the ketogenic diet (defined as any diet containing <50 grams of carbohydrates/day^[34]^) indicates a negative/adverse link (Model 3). The high-protein diet (defined as a daily protein intake of >1.5 grams of protein per kilogram of body weight^[35]^) exhibits beneficial links with increased grey and white matter volumes, though undesirably associated with increased phenotypic age.

Table 2, panel C, further reveals nutrient-level associations with aging outcomes. In particular, carbohydrate intake is positively/beneficially linked with all aging measures, conditional on protein and fat intakes (Models 1-4). On the other hand, protein and fat intakes are associated with reduced grey matter volume, and fat intake is further linked with increased phenotypic age and reduced white matter volume.

Table 2, panel D, reveals beneficial associations between diet quality and aging outcomes. Higher scores of HEI-2015 and IDDS are both beneficially linked with increased telomere length (Model 1) and reduced phenotypic age (Model 2). In addition, a higher DASH is linked with reduced phenotypic age. No significant associations between diet-quality scores and grey/white matter volumes are found.

The HDFE estimates presented above provide initial evidence that many dietary habits, dietary patterns, and macronutrient intakes are closely connected with aging, albeit in mixed directions. The chord diagram in Figure 1 presents all statistically significant estimates (*p* <0.05) of dietary factors on four measures of aging, marking positive/beneficial links in green and negative/adverse links in red. Overall, these findings suggest that a healthy-aging diet may contain reduced processed meat and red meat (e.g., beef and lamb) and increased cereal/bread, fruits/vegetables, and white meat (e.g., non-oily fish).

**Figure 1.**
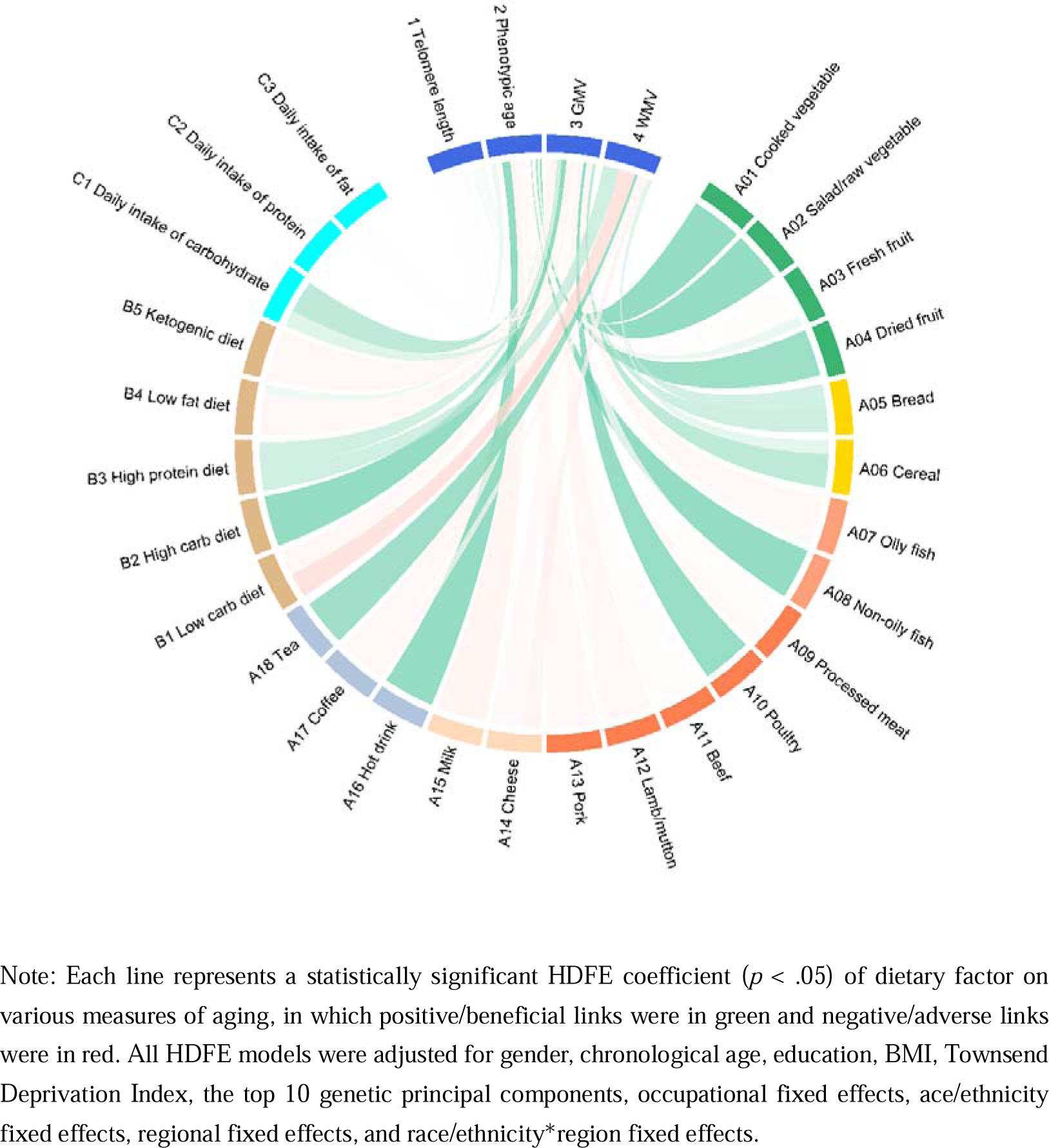
Chord diagram of associations of dietary factors with aging.

### Causal links between macronutrient intakes and aging

To address unobserved confounding that may contaminate the estimated food-intake and dietary-habit effects on aging, we employ the MVMR method to identify the causal effects of three macronutrient intakes simultaneously.^[28,31,36]^ As noted above, MVMR uses genetic variants associated with diet compositions of carbohydrates, protein, and fat as IVs for their intakes. Nine independent SNPs (see section 2.5) were selected as genetic IVs based on recent GWAS findings.^[30]^ The validity of jointly using these SNPs as genetic IVs has been established.^[31]^ We further control for the top 10 genetic PCs and occupational, ethnic, and regional FEs in all MVMR models.

As shown in Table 3, the first-stage F-statistics from all MVMR models exceeded the conventional cut-off of 10 for weak-IV tests, suggesting the nine SNPs jointly as strong IVs in our MVMR design.^2^ Meanwhile, overidentification (Sargan) tests revealed no evidence of directional pleiotropy, suggesting that our genetic IVs were not correlated with unobserved confounders.

**Table 3.**
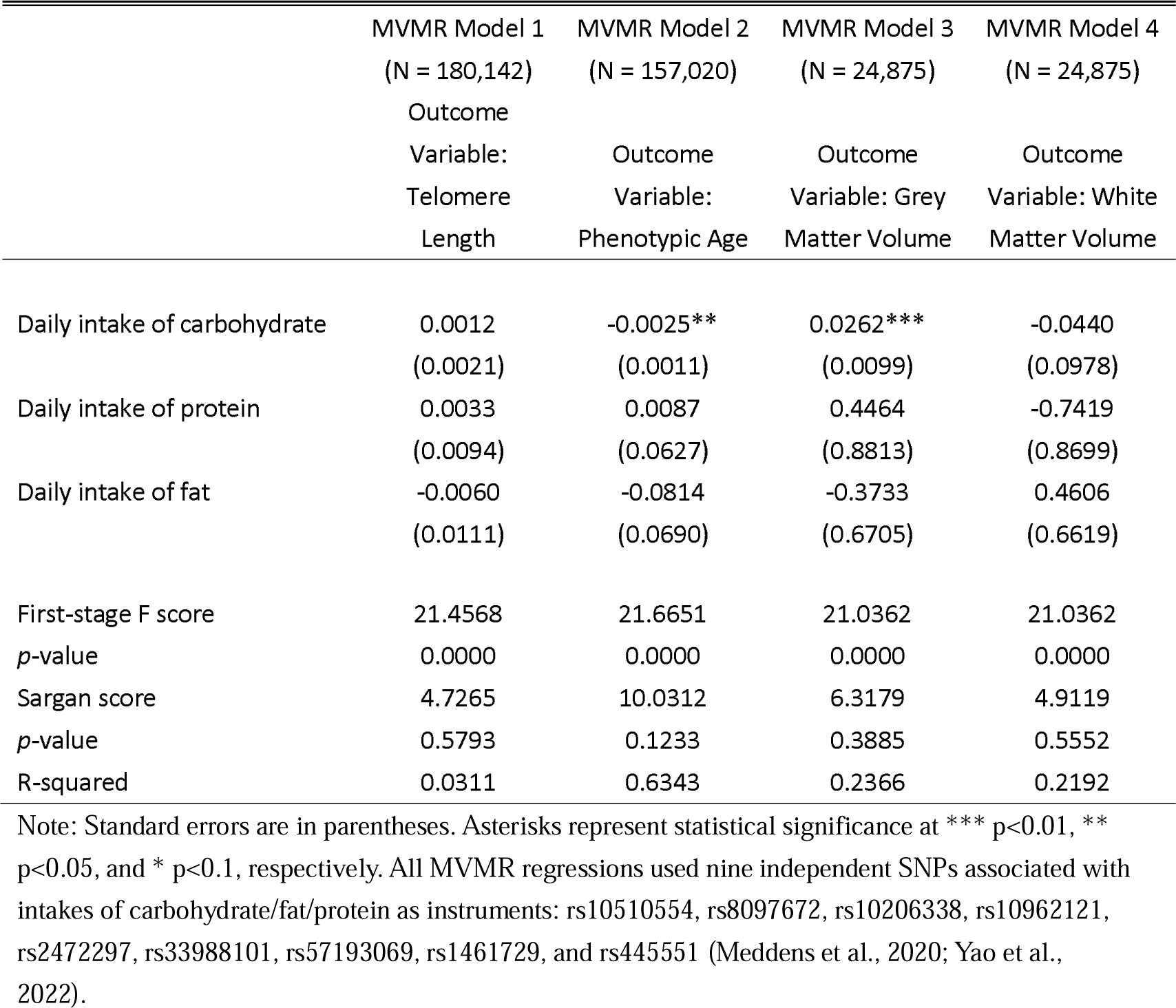
Causal associations of macronutrients with aging from Multivariable Mendelian Randomization (MVMR) estimation.

The MVMR estimates (Table 3) suggest a potential protective effect of carbohydrate intake against aging, in terms of both reduced phenotypic age (Model 2) and increased grey matter volume (Model 3). For example, a 10-gram increment in daily carbohydrate intake is associated with a 0.025-year (95% CI=[-0.047, -0.003], *p*=0.026) reduction in phenotypic age and a 0.26-cm^3^ (95% CI=[0.07, 0.46], *p*=0.008) increase in grey matter volume. In contrast, MVMR estimates for protein and fat intakes reveal no significant impacts on aging.

To further assess potential nonlinear effects of carbohydrate intake on aging, we performed a semiparametric nonlinear Mendelian Randomization (NLMR) estimation.^[37,38]^ Estimates from NLMR confirmed the beneficial linear effects of carbohydrate intake on aging but detected no significant nonlinear effect of carbohydrate intake (nonlinearity *p*-value=0.386).

## Discussion

This study identified several dietary factors that are significantly linked with the acceleration or slowdown of aging; in particular, the protective effect of carbohydrate intake against aging is causal and robust. These findings emphasize the role diet plays in delaying or speeding the biological aging process, highlighting the potential of a carbohydrate-rich diet in promoting healthy aging.

Previous studies have reported that carbohydrate-rich diets were associated with accelerated aging in model organisms, such as yeast and roundworms (*Caenorhabditis Elegans*). However, direct evidence regarding whether different levels of carbohydrate intake influence aging in humans remains lacking.^[39,40]^ An RCT involving 63 obese persons found that a low-carbohydrate diet caused significant weight loss.^[41]^ Our results, however, suggest that a low-carbohydrate diet may adversely impact aging despite its weight-reduction effect and related effects of reducing risks of coronary heart diseases.

Previous studies documented the associations of calorie restriction with increased longevity and improved health in both model organisms and humans.^[42,43]^ To examine the role of caloric intake, we ran additional HDFE regressions of different aging measures on daily total caloric intake (in 1,000 calories). The first set of HDFE models only controlled for the set of FEs previously described; the second set of models further controlled for individuals’ gender, chronological age, education, BMI, and TDI.^3^ As reported in Supplementary eTable 4, the associations of caloric intake with aging outcomes were *not* robust once conditional on the set of individual factors. Hence, consistent with previous findings,^[44]^ our findings suggest that nutritional composition in diets, rather than caloric restriction, is what matters for aging.

Our findings have multiple potential explanations. First, phenotypic age used in our study is an epigenetic biomarker of aging that captures morbidity and mortality risks^[45]^, and a low-carbohydrate diet (<40%) conferred the greatest mortality risk.^[46]^ Second, Jensen et al. (2021)^[47]^ reported that diets rich in fruits and vegetables and low in meat are associated with increased brain volume and connectivity. The *CPLX3* gene, a prominent marker of subplate neurons in the brain that regulate cortical development and plasticity, is also associated with cereal intake.^[48]^ In our sample, higher daily carbohydrate intake is correlated with higher intakes of cereal, bread, and fresh/dried fruits, while a low-carbohydrate diet is correlated with higher intakes of lamb/mutton, beef, and processed meats (Supplementary eFigure 1). More recently, Yao et al. (2022)^[31]^ reported a causal relationship between higher relative carbohydrate intake and lower depression risk, in which depression and cognitive dysfunction are considered to share a common neuropathological platform in cortical and sub-cortical brain areas.^[49]^

Several limitations should be noted when interpreting our results. First, the participants were dominantly white, relatively healthy, and well-educated, which may limit the generalizability of our findings to other populations. Second, MVMR estimates from unrelated individuals can be biased due to population structure, assortative mating, and genetic nurture. Future research may circumvent such biases by employing a within-family design. Finally, our study does not provide an understanding of the biological mechanism underlying the beneficial effect of carbohydrate intake on aging. Illumination of the causal mechanism may require randomized diet interventions in the future.

### Conclusions

Our findings demonstrate that diet plays a key role in the biological aging process, suggesting that a healthy-aging diet may include reduced processed/red meat consumption and increased cereal/bread, fruit/vegetables, and white meat intakes. Carbohydrate intake has a protective effect on aging. This finding can help inform the design and implementation of dietary interventions to promote healthy aging.

## Data Availability

The GWAS summary statistics used were available from the MRC-IEU OpenGWAS project. Data from UK Biobank are available to all researchers upon making an application. This research has been conducted using the UK Biobank Resource under Applications 89068.

## Methods

### Study sample and data

Our analysis draws dietary, biomarker, and socio-demographic data from the UK Biobank baseline survey. UK Biobank is the database for a population-based study involving 502,409 UK residents approved by the NHS National Research Ethics Service (Ref: 11/NW/0382). Between March 2006 and July 2010, individuals residing within 25 miles of one of the 22 study assessment centers in England, Scotland, and Wales were recruited to provide data on a wide range of socio-demographic, clinical, and lifestyle outcomes. Blood, urine, and saliva samples, as well as physical measurements, were collected from all participants with their written informed consent during the interviews.

At the baseline (2006), a food frequency touchscreen questionnaire was used to collect data on all participants’ dietary habits,^[15]^ including single food intakes (e.g., cooked vegetables consumed), food types (e.g., types of cereal primarily consumed), and intake frequencies (e.g., frequency of oily fish consumption). Observations (7,848 participants) with unrealistic answers were dropped. In early 2009, the main UK Biobank study protocol further included the Oxford WebQ, a web-based dietary assessment tool that asks about the consumption of up to 206 types of foods and 32 types of drinks during the previous 24 hours.^[16,17]^ In total, 210,977 participants had completed at least one assessment.

Serum biomarker data used to construct phenotypic age were obtained from the UK Biobank blood samples collected from all participants at the baseline and analyzed within 24 hours of the blood draw with Beckman Coulter LH750 instruments at the UK Biobank central laboratory.^[18]^

Participants’ genotypes were analyzed with the Affymetrix (Santa Clara, CA, USA) UK Biobank Axiom Array and the UK BiLEVE Axiom Array. Information regarding principal components analysis and genetic principal components is provided elsewhere.^[19]^ Telomere length data measuring biological age were also derived from the DNA samples.^[10]^

High-quality MRI (magnetic resonance imaging) brain imaging data were available for a subset of participants (*N*=42,942) upon additional agreement. These data were acquired using a Siemens Skyra 3T scanner (Siemens Healthcare, Erlangen, Germany) with a standard 32-channel head coil. Structural imaging and diffusion data were processed by UK Biobank technicians and made available to approved researchers as imaging-derived phenotypes (IDPs).^[21,22]^

### Measurements of dietary factors

The consumption of seven common food groups (vegetables, fruits, grains, fish, meat, dairy products, and beverages) and intakes of 18 specific food categories were investigated:^4^cooked vegetables (tablespoons/day), salad/raw vegetables (tablespoons/day), fresh fruits (pieces/day), dried fruits (pieces/day), bread (pieces/day), cereal (bowls/week), oily fish (times/week), non-oily fish (times/week), processed meat (times/week), poultry (times/week), beef (times/week), lamb/mutton (times/week), pork (times/week); cheese (times/week), milk (full cream/semi-skimmed/skimmed=1; never/rarely consumed/soya=0), hot drinks (very hot/hot=1; warm/never hot drinks=0), coffee (cups/day), and tea (cups/day).

#### Macronutrient intake

Daily macronutrient intakes of carbohydrates, protein, and fat (all in grams) were calculated by UK Biobank experts based on responses from the computerized 24-hour dietary recall follow-up questionnaire (described above).^[16,17]^ Data are available for 210,977 participants in this study.

#### Dietary patterns

This study considers six dietary patterns: low-carb, balanced-carb, high-carb, high-protein, low-fat, and ketogenic diets, which are defined based on the absolute intake of a macronutrient as a function of body weight or a proportion to total energy intake—Supplemental eTable 2 provides more details.

#### Diet quality

Following detailed protocols provided by FAO (Food and Agriculture Organization) and USDA (US Department of Agriculture) guidelines, we calculated three diet-quality scores using the 24-hour dietary recall data: individual Diet Diversity Score (IDDS),^[23, 24]^ Healthy Eating index (HEI-2015),^[25]^ and Dietary Approaches to Stop Hypertension (DASH) index.^[26]^

### Measurements of aging

#### Telomere length

Leucocyte telomere length (LTL) was measured using an established multiplex qPCR assay on DNA samples from UK Biobank participants. After extensive quality checks and adjustments for technical factors, 473,994 participants in our sample had valid LTL measurements. The LTL was then log-transformed to yield a normal distribution and z-standardized using the distribution of all individuals with valid LTL measurements.^[20]^

#### Phenotypic age

Phenotypic age was constructed using chronological age and nine clinical biomarkers (albumin, creatinine, glucose, C-reactive protein, lymphocyte percentage, mean corpuscular volume, red cell distribution width, alkaline phosphatase, and white blood cell count) in a parametric proportional hazards model based on the Gompertz distribution.^[4]^ The biomarker data were obtained from UK Biobank biological samples collected upon participant enrollment, typically analyzed at the UK Biobank central laboratory within 24 hours of the blood draw with Beckman Coulter LH750 instruments.

#### Grey/white matter volume

Participants’ whole-brain grey and white matter volume data (normalized for head size) were obtained from the UK Biobank brain imaging datasets.

### Covariates

To control for potential confounding factors, we included a series of socio-demographic, physiological, and genetic characteristics in the statistical analysis: gender, chronological age, education (whether with a college/university degree), body mass index (BMI), Townsend Deprivation Index (TDI),^5^ and top ten genetic principal components (PCs).

### Statistical models

We used four measures of aging (telomere length, phenotypic age, whole-brain grey matter volume, and whole-brain white matter volume) as the outcomes and four sets of dietary factors (dietary habits/food intake, dietary patterns, macronutrient intakes, and diet-quality scores) as main explanatory variables.

First, high-dimensional fixed effects (HDFE) models^[27]^ were fitted to examine the associations of dietary factors with aging while controlling for a large set of (occupational, ethnicity, regional, and ethnicity-by-region) fixed effects (FEs) to mitigate unobserved confounding. The models take the following form:

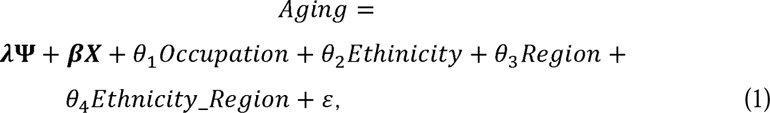

where *Aging* is one of the four aging measures of interest; Ψ represents a particular set of dietary factors (e.g., dietary patterns or macronutrient intakes); λ is the corresponding parameter vector; ***X*** is the set of covariates discussed above; also included are a total of 477 FEs: 336 occupational, 18 ethnicity, 10 regional, and 113 ethnicity-by-region FEs; ε is a disturbance term.

Second, since macronutrient intakes are potentially endogenously chosen, we employ a Multivariate Mendelian Randomization (MVMR) framework to address this concern. To facilitate causal identification, MVMR uses genetic variants as instrumental variables (IV) to simultaneously identify the effects of multiple endogenous exposures on an outcome.^[28]^ The strength of Mendelian Randomization stems from the random allocation of genes at meiosis, resembling random treatment assignments in RCTs that may be infeasible or unethical in our setting.^[29]^ Based on recent genome-wide association studies (GWAS) identifying genetic variants that are closely related to diet compositions and macronutrient intakes,^[30]^ we selected nine independent single nucleotide polymorphisms (SNPs)—rs10510554, rs8097672, rs10206338, rs10962121, rs2472297, rs33988101, rs57193069, rs1461729, and rs445551—as genetic IVs in our MVMR design.^[31]^ Formally, we estimate the following two-stage least-squares (2SLS) models:

#### First stage

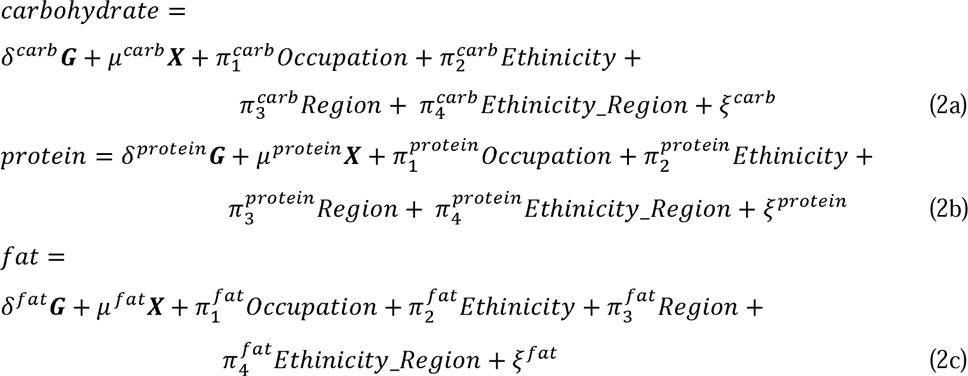

#### Second stage

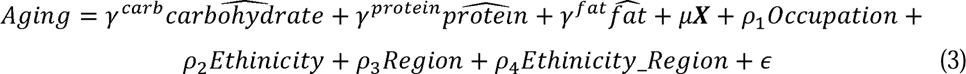

Mechanically, in the first stage (equations 2a-2c), each macronutrient intake is regressed on the nine genetic IVs (G), the set of covariates (***X***), and the full set of FEs described earlier. In the second stage (equation 3), each aging outcome is regressed on the fitted values of all three macronutrient intakes simultaneously, along with all covariates used in the first stage. In practice, all regressions involved in the 2SLS framework were performed at once using STATA 15.

## Footnotes

1 Interestingly, poultry intake is associated with increased brain grey/white matter volumes.

2 Supplementary eTable 3 reports detailed first-stage regression results.

3 Due to high correlations between daily intakes of calories and carbohydrate (correlation coefficient ρ = 0.8591), fat (ρ = 0.8511), and protein (ρ = 0.75), we did not directly include daily caloric intake in the main models to avoid multicollinearity.

4 Supplemental eTable 1 provides detailed data field codes for variable constructions.

5 The TDI Index is a measure of local material deprivation: positive (negative) numbers indicate lower (higher) socioeconomic status.

## Notes

### Competing Interest Statement

The authors have declared no competing interest.

### Funding Statement

This research was funded by the National Natural Science Foundation of China (Grant No. 72103187) and the 2115 Talent Development Program of China Agricultural University.

### Author Declarations

The study was approved by the North West Multicenter Research Ethics Committee (REC reference for UK Biobank 11/NW/0382), and all participants provided written informed consent.

