## Supplementary Tables and Figures for "Do dietary habits, dietary patterns, and macronutrient intakes affect aging? Multi-dimensional evidence from UK Biobank"

### Supplementary Materials

**Contents:**

**Supplementary eTable 1-4**

**Supplementary eFigure 1**

**Supplementary References**

Supplementary eTable 1. Detailed UKB data field codes used in the study

| Category | Variable Description | UKB Data Field Code |
|---|---|---|
| Dietary habits/food intake | Cooked vegetable intake | 1289 |
| Dietary habits/food intake | Salad / raw vegetable intake | 1299 |
| Dietary habits/food intake | Fresh fruit intake | 1309 |
| Dietary habits/food intake | Dried fruit intake | 1319 |
| Dietary habits/food intake | Bread intake | 1438 |
| Dietary habits/food intake | Cereal intake | 1458 |
| Dietary habits/food intake | Oily fish intake[a] | 1329 |
| Dietary habits/food intake | Non-oily fish intake[b] | 1339 |
| Dietary habits/food intake | Processed meat intake | 1349 |
| Dietary habits/food intake | Poultry intake | 1359 |
| Dietary habits/food intake | Beef intake | 1369 |
| Dietary habits/food intake | Lamb/mutton intake | 1379 |
| Dietary habits/food intake | Pork intake | 1389 |
| Dietary habits/food intake | Cheese intake | 1408 |
| Dietary habits/food intake | Milk type used | 1418 |
| Dietary habits/food intake | Hot drink temperature | 1518 |
| Dietary habits/food intake | Coffee intake | 1498 |
| Dietary habits/food intake | Tea intake | 1488 |
| Macronutrient intake | Carbohydrate | 26013 |
| Macronutrient intake | Protein | 26005 |
| Macronutrient intake | Fat | 26008 |
| Telomere length | Z-adjusted T/S log | 22192 |
| Phenotypic age | Albumin | 30600 |
| Phenotypic age | Creatinine | 30700 |
| Phenotypic age | Glucose | 30740 |
| Phenotypic age | C-reactive protein | 30710 |
| Phenotypic age | Lymphocyte percentage | 30180 |
| Phenotypic age | Mean corpuscular volume | 30040 |
| Phenotypic age | Red blood cell (erythrocyte) distribution width | 30070 |
| Phenotypic age | Alkaline phosphatase | 30610 |
| Phenotypic age | White blood cell (leukocyte) count | 30000 |
| Grey/white matter volume | Volume of grey matter (normalised for head size) | 25005 |
| Grey/white matter volume | Volume of white matter (normalised for head size) | 25007 |

Note: [a.] In UK Biobank, oily fish includes: salmon, anchovies, trout, swordfish, mackerel, bloater, herring, cacha, sardines, carp, pilchards, hilsa, kipper, jack fish, eel, katla, whitebait, orange roughy, tuna (fresh only), pangas, and sprats (https://biobank.ndph.ox.ac.uk/showcase/field.cgi?id=1329). [b.] In UK Biobank, non-oily fish (or white fish) includes: cod, tinned tuna, and haddock (https://biobank.ndph.ox.ac.uk/showcase/field.cgi?id=1339).

Supplementary eTable 2. Definitions and sources of dietary patterns

| Dietary patterns | Definition | Source |
|---|---|---|
| Low-carb diet | Defined as any diet containing carbohydrates less than 45% of the daily total energy intake. | Naude et al. (2014) |
| Balanced-carb diet | Defined as any diet containing carbohydrates between 45% to 65% of the daily total energy intake. | Naude et al. (2014) |
| High-carb diet | Defined as any diet containing carbohydrates of more than 65% of the daily total energy intake. | Naude et al. (2014) |
| High-protein diet | Defined as a daily protein intake of more than 1.5 grams of protein per kilogram of body weight. | Ko et al. (2020) |
| Low-fat diet | Defined as any diet containing fat less than 30% of the daily total energy intake. | Yancy et al. (2004) |
| Ketogenic diet | Defined as any diet containing less than 50 grams of carbohydrates a day. | Bueno et al. (2013) |

Supplementary eTable 3. First stage regression results of MVMR

| VARIABLES | (1)<br>Daily intake of carbohydrate | (2)<br>Daily intake of protein | (3)<br>Daily intake of fat |
|---|---|---|---|
| rs33988101 | 0.8650*** | -0.4431*** | -0.4520*** |
| | (0.2894) | (0.0839) | (0.0993) |
| rs10962121 | 0.5028* | -0.3111*** | -0.1712* |
| | (0.2866) | (0.0831) | (0.0984) |
| rs10510554 | 0.3345 | -0.3745*** | -0.3376*** |
| | (0.2889) | (0.0838) | (0.0992) |
| rs8097672 | 1.2844*** | -0.1518 | -0.0702 |
| | (0.4083) | (0.1185) | (0.1402) |
| rs10206338 | -0.8909*** | -0.0140 | 0.0897 |
| | (0.2894) | (0.0840) | (0.0993) |
| rs2472297 | -0.8410** | 0.2422** | 0.2687** |
| | (0.3300) | (0.0957) | (0.1133) |
| rs57193069 | 0.4099 | 0.0307 | -0.1272 |
| | (0.2869) | (0.0832) | (0.0985) |
| rs1461729 | -1.8420*** | 0.2032 | -0.0301 |
| | (0.4737) | (0.1374) | (0.1626) |
| rs445551 | -0.2499 | 0.3127*** | 0.0918 |
| | (0.3086) | (0.0895) | (0.1059) |
| Male | 34.2084*** | 8.7068*** | 10.9569*** |
| | (0.4111) | (0.1193) | (0.1411) |
| College | 2.9467*** | 0.5379*** | 1.4210*** |
| | (0.4206) | (0.1220) | (0.1444) |
| Age | -0.0989*** | -0.0772*** | -0.1815*** |
| | (0.0260) | (0.0076) | (0.0089) |
| BMI | -0.9033*** | 0.3534*** | 0.0930*** |
| | (0.0484) | (0.0140) | (0.0166) |
| Townsend deprivation index | 0.2826*** | -0.2149*** | 0.1584*** |
| | (0.0768) | (0.0223) | (0.0264) |
| Constant | 265.7449*** | 73.8014*** | 80.4121*** |
| | (2.2630) | (0.6565) | (0.7767) |
| Top 10 PCs | Y | Y | Y |
| Occupational FE | Y | Y | Y |
| Ethnicity FE | Y | Y | Y |
| Regional FE | Y | Y | Y |
| Observations | 180,142 | 180,142 | 180,142 |
| R-squared | 0.0396 | 0.0377 | 0.0384 |

Supplementary eTable 4. Test for association between caloric intake and aging

| | (A) HDFE Set 1 | | | | (B) HDFE Set 2 | | | |
|---|---|---|---|---|---|---|---|---|
| | (1) | (2) | (3) | (4) | (5) | (6) | (7) | (8) |
| VARIABLES | Outcome Variable: Telomere Length | Outcome Variable: Phenotypic Age | Outcome Variable: Grey Matter Volume | Outcome Variable: White Matter Volume | Outcome Variable: Telomere Length | Outcome Variable: Phenotypic Age | Outcome Variable: Grey Matter Volume | Outcome Variable: White Matter Volume |
| Daily intake of calories (in kilocalories) | -0.0186*** | 0.2589*** | -4.9597*** | 1.4989*** | 0.0016 | -0.0233 | -1.0731 | -0.3232 |
| | (0.0035) | (0.0328) | (0.4730) | (0.4143) | (0.0035) | (0.0154) | (1.3793) | (0.4111) |
| Male | - | - | - | - | -0.1721*** | 3.3731*** | -24.3023*** | 9.2045*** |
| | | | | | (0.0053) | (0.0229) | (0.5299) | (0.5743) |
| College | - | - | - | - | 0.0378*** | -0.3519*** | -2.5499*** | -1.5360*** |
| | | | | | (0.0051) | (0.0223) | (0.5044) | (0.5467) |
| Age | - | - | - | - | -0.0223*** | 0.9858*** | -3.6492*** | -1.6191*** |
| | | | | | (0.0003) | (0.0013) | (0.0326) | (0.0354) |
| BMI | - | - | - | - | -0.0056*** | 0.3087*** | -1.0244*** | 0.0859 |
| | | | | | (0.0005) | (0.0023) | (0.0566) | (0.0613) |
| Townsend deprivation index | - | - | - | - | -0.0015* | 0.0361*** | -0.2675*** | -0.1075 |
| | | | | | (0.0008) | (0.0037) | (0.0883) | (0.0957) |
| Constant | 0.0827*** | 18.6919*** | 801.8828*** | 697.3982*** | 1.4963*** | -45.5426*** | 1,033.8103*** | 784.3823*** |
| | (0.0078) | (0.0731) | (1.0776) | (0.9439) | (0.0232) | (0.1014) | (2.4568) | (2.6626) |
| Top 10 PCs | Y | Y | Y | Y | Y | Y | Y | Y |
| Occupational FE | Y | Y | Y | Y | Y | Y | Y | Y |
| Ethnicity FE | Y | Y | Y | Y | Y | Y | Y | Y |
| Regional FE | Y | Y | Y | Y | Y | Y | Y | Y |
| Ethnicity*Region FE | Y | Y | Y | Y | Y | Y | Y | Y |
| Observations | 196,198 | 170,890 | 26,925 | 26,925 | 195,951 | 170,681 | 26,900 | 26,900 |
| R-squared | 0.0179 | 0.1386 | 0.1042 | 0.0524 | 0.0542 | 0.8198 | 0.4593 | 0.1252 |

Note: Standard errors are in parentheses. Asterisks represent statistical significance at *** p<0.01, ** p<0.05, and * p<0.1, respectively.

Supplementary eFigure 1. Correlation coefficients of dietary factors

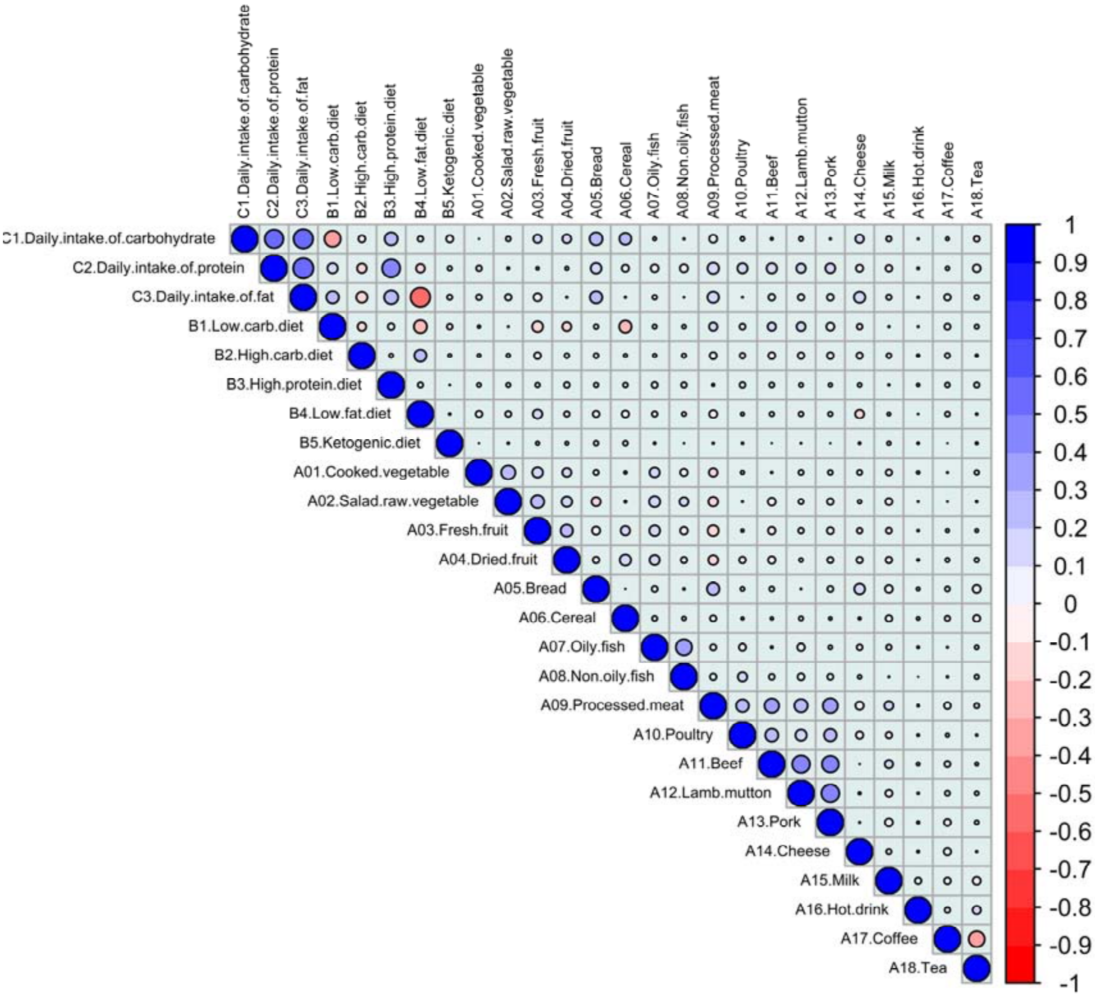
